# A prospective double-blinded non-selection trial of reproductive outcomes and chromosomal normalcy of newborns derived from putative low/moderate-degree mosaic IVF embryos

**DOI:** 10.1101/2021.02.07.21251201

**Authors:** A. Capalbo, M. Poli, L. Rienzi, L. Girardi, D. Cimadomo, F. Benini, A. Farcomeni, J. Cuzzi, C. Rubio, E. Albani, L. Sacchi, A. Vaiarelli, I. Vogel, E. Hoffmann, C. Livi, P.E. Levi-Setti, F.M. Ubaldi, C. Simón

## Abstract

**Background:** Next generation sequencing (NGS) has increased detection sensitivity of intermediate chromosome copy number variations (CNV) consistent with chromosomal mosaicism. Recently, this methodology has found application in preimplantation genetic testing (PGT) of trophectoderm (TE) biopsies collected from IVF-generated human embryos. As a consequence, the detection rate of intermediate CNV states in IVF embryos has drastically increased, posing questions about the accuracy in identifying genuine mosaicism in highly heterogeneous biological specimens. The association between analytical values consistent with mosaicism and the reproductive potential of the embryo, as well as newborn’s chromosomal normalcy, have not yet been thoroughly determined.

**Methods:** We conducted a multicentre, double-blinded, non-selection trial including 1,190 patients undergoing in a total of 1,337 IVF with preimplantation genetic testing for aneuploidies (PGT-A) treatment cycles. NGS was performed on clinical TE biopsies collected from blastocyst-stage embryos. All embryos were reported as euploid if all autosomes had a chromosomal copy number value below the threshold of 50% abnormal cells per sample. After embryo transfer, three comparative classes were analysed: uniformly euploid profiles (<20% aneuploid cells), putative low-degree mosaicism (20%-30% aneuploid cells) or putative moderate-degree mosaicism (30%-50% aneuploid cells). Primary outcome measure was live birth rate (LBR) per transfer and newborn’s karyotype.

**Results:** LBR after transfer of uniformly euploid embryos, low-degree, and moderate-degree mosaic embryos were 43.4% (95% C.I. 38.9 - 47.9), 42.9% (95% C.I. 37.1 - 48.9) and 42% (95% C.I. 33.4 - 50.9), respectively. No difference was detected for this primary outcome between euploid and mosaic low/moderate categories (OR= 0.96; 95% CI 0.743 to 1.263; P=0.816). The non-inferiority endpoint was met as the confidence interval for the difference fell below the planned 7.5% margin (95% C.I. −5.7 - 7.3). Likewise, no statistically significant difference was observed comparing moderate versus low degree mosaic embryos (P=0.92). Neonatal karyotypes were also similar and no instances of mosaicism or uniparental disomies (UPDs) were detected in babies born following putative low or moderate-degree mosaic embryo transfer. Should the embryos with low or moderate-degree mosaic TE biopsies had been classified as chromosomally abnormal and thus discarded for clinical use, LBR per cycle would have decreased by 36% without any clinical benefit.

**Conclusions:** This prospective non-selection trial provides substantial evidence that reporting and/or not transferring embryos with low/moderate-degree mosaicism for whole chromosomes have no clinical utility. Moreover, dismissing these embryos from clinical use has the counterproductive effect of reducing overall embryo availability, thus reducing the chance of successful outcome derived from an IVF treatment without any clinical benefit. (Funded by Igenomix; ClinicalTrials.gov number, NCT03673592)

## INTRODUCTION

Mosaicism is defined as the presence of two or more genotypically different cell lines in a given organism, embryo, or cell line. Although its presence has been documented in less than 0.5% of prenatal specimens (e.g., detected through amniocentesis) and no evidence of increased incidence has been shown in babies born following IVF treatments ^1,2^, chromosomal mosaicism has been reported in up to 73% of cleavage stage human preimplantation embryos ^3^. Due to this alleged high incidence, combined with improved analytical sensibility provided by next generation sequencing (NGS), mosaicism has recently attracted the attention of the scientific community in relation to its impact on embryo viability and reproductive outcome of *in vitro* fertilization (IVF) cycles with preimplantation genetic testing for aneuploidies (PGT-A) ^4^.

Diagnosing embryo mosaicism based on the detection of altered chromosomal profiles in a single trophectoderm biopsy is challenged by several biological and technical considerations ^5^. Moreover, the reporting of embryo mosaicism diagnosis generates a course of action that often has major impact on clinical practice. A recent web-based survey showed that almost 50% of IVF centers performing PGT-A consider an embryo as mosaic when abnormal cells are expected to be present in ≥20% of the tested sample ^6^. The same survey also reported that only around 30% of patients with a result consistent with mosaicism would accept the transfer of the embryo after a genetic counseling session. Because of the unknown clinical impact of embryo mosaicism on either IVF outcome (e.g., pregnancy and miscarriage rates) and ensuing offspring (e.g., chromosomal abnormalities), uniformly euploid embryos are commonly prioritized for transfer to the patient, whilst putative mosaic ones are given low priority or even discarded ^7–9^. Evidence from actual clinical data indicate that fewer than 3% of embryos with a putative mosaic diagnosis are selected for clinical use ^7^.

Presently, clinical outcomes of putative mosaic embryos have only been compared retrospectively using embryos analysed with prior technologies (e.g., array-comparative genomic hybridization)^4^ and in selected subpopulations of patients that failed to get pregnant with previous euploid embryos ^7,10–12^. Inherently, past studies introduced a strong bias in the evaluation of clinical outcomes as putative mosaic embryos were mainly employed in patients with poorer prognosis, demonstrated by previous failed implantations following the transfer of one or several euploid embryos.

This NGS-based prospective non-selection study was designed to allow an unbiased assessment of the reproductive potential and offspring chromosomal normalcy between uniformly euploid and putative mosaic embryos, providing definitive evidence on their clinical performance. Here, we report that putative mosaic embryos show similar clinical outcome to uniformly euploid embryos, without any significant implication for pregnancy and live birth outcomes or the offspring’s chromosomal health.

## METHODS

### Study design and participants

We conducted a multicentre, blinded, non-selection trial involving consecutive patients who underwent IVF with blastocyst stage preimplantation genetic testing for aneuploidies (PGT-A) followed by single frozen euploid, low, or moderate-degree mosaic embryo transfer (SET). Patients below the age of 45, using autologous oocytes, undergoing ICSI for all oocytes and had at least one transferrable embryo available (euploid or low/moderate-grade mosaic) were eligible for participation to this study. Patients were excluded from the study if no embryo was suitable for biopsy, if the embryo to be transferred showed the worst morphological grade according to an adaptation of Gardner’s criteria ^13^ or if the female patient had a chronic medical condition associated with adverse pregnancy outcomes (see **Supplementary Appendix** for a detailed description of exclusion and inclusion criteria). Ovarian stimulation, embryo culture system, embryo biopsy technique and luteal-phase support were carried out according to standard practices employed at each clinic (see **Supplementary Appendix** for a detailed description of methods employed).

The trial was conducted in compliance with the International Conference on Harmonisation and the Declaration of Helsinki. The protocol was approved by the Institutional Review Board of Clinica Valle Giulia, Rome (3 September 2018) and Humanitas Research Hospital Ethics Committee, Rozzano (code 477/19). All the patients provided written informed consent before participation. The trial was supported by Igenomix and registered to ClinicalTrials.gov as NCT03673592.

### Intervention

Following TE biopsy and NGS-based chromosomal analysis, a diagnostic report on the embryos’ chromosomal status was sent to the clinical sites (see **Supplementary Appendix** for the complete PGT-A protocol). Embryos showing low or moderate degree of chromosomal mosaicism were blindly reported as euploid without distinction from uniformly euploid embryos. Among those reported as euploid, embryos were selected for transfer based on standard morphological features, providing blinded allocation of patients into the three main categories “Euploid” Group A, “Low-degree mosaic” Group B (20%-30% of aneuploid cells), and “Moderate-degree mosaic” Group C (30%-50% of aneuploid cells). Cases were followed-up during the post-transfer, gestational and post-natal periods. The chromosomal status of 38 newborns derived from the transfer of putative mosaic embryos was investigated using single nucleotide polymorphism arrays (SNPa genotyping) on saliva samples collected from the newborns and their parents. Genotyping data of the trios was used to investigate any potential instance of mosaicism or uniparental disomies (UPD) in the offspring ensued following putative mosaic SET (see **Supplementary Appendix** for details of the genotyping protocol).

### Outcomes

Primary outcomes were live birth rate (LBR) per transferred embryo, defined as the live birth of a newborn delivered on or after 24 weeks of gestation over the number of embryos replaced, and newborn’s karyotype. Secondary outcomes were pregnancy rate (PR), implantation rate (IR), biochemical pregnancy (BP), and clinical miscarriage (CM). Additional secondary outcomes included mean gestational age at birth and birth weight (see **Supplementary Appendix** for the complete outcomes description). Adverse outcome included the detection of chromosomal abnormalities (whether in uniform or mosaic conformation as well as uniparental disomy) in miscarried product of conception (POC), during prenatal diagnosis (PND; amniocentesis/chorionic villi sampling, CVS) and/or at birth. Definitions of the secondary efficacy and safety outcomes are provided in the **Supplementary Appendix**.

The implication of excluding putative mosaic embryos from clinical use has been evaluated in consideration of the potential loss of live births in a given IVF treatment cycle (CLBR per cycle). The cumulative LBR for a complete cycle is defined as the chance of a live birth from an ovarian stimulation cycle including all subsequent FETs from that cycle ^14,15^. Two scenarios were analysed: i) using actual data from embryo transfer in the study period excluding live births achieved from low and moderate mosaic embryos; ii) by modeling the optimistic scenario where all transferable embryos are replaced. For this second approach, a probabilistic projection was computed accounting for all euploid embryos with or without putative mosaic embryos produced from a single ovarian stimulation cycle and considering the combined probability of achieving a live birth based on the available embryos. In this model, the CLBR per cycle was computed by an optimistic approach, that is assuming that all available embryos are transferred in a given cycle and with a defined probability of success. Live birth rate per euploid or mosaic embryo was the actual value observed in the study across the three study groups (i.e., 43%). The observed and projected CLBR per cycle analysis (with and without the clinical use of putative mosaic embryos) is shown across all female ages.

### Statistical analysis

The primary endpoint for this analysis was non-inferiority of LBR when comparing euploid vs mosaic embryos. Assuming a LBR of 45% for uniformly euploid embryos vs 42.5% for moderate or low degree mosaics ^16^, assuming a 1:1 sampling ratio for the two groups, and a planning non-inferiority margin of 7.5% ^17,18^, we calculated that 421 embryos per group would guarantee a power of at least 90% for a significance level fixed at 5%. This sample size was also >90% powered to claim non-inferiority in the miscarriage rate between control and test group with a margin of 2%, assuming a 10% rate for uniformly euploid embryos and 15% for moderate/low mosaics. Data are expressed as mean +/- standard deviation or percentages as appropriate. Proportions were compared using chi-squared test, or Fisher exact test for 2 x 2 contingency tables. The non-inferiority endpoint was set as the 95% C.I. for difference in proportions laying below the planned margin. In addition to computing confidence intervals and p-values for difference in proportions, we computed the odds-ratios (OR) and adjusted odds-ratios (AOR) of mosaicism for LBR, PR, IR, MC, and BP through logistic regression models. In multivariate analyses, odds-ratios were adjusted for female age, male age, centre, morphology of the blastocyst, day of biopsy, number of previous implantation failures, previous miscarriages, previous live birth, indication, and sperm origin (ejaculated vs surgical). All tests were two-tailed. All analyses were conducted using SPSS v. 21 and R v. 3.5.1.

### Single nucleotide polymorphism (SNP) analysis of aneuploidies and uniparental disomy

We used the Human CytoSNP-12 to genotype 293, 552 SNPs genomewide in parents and the proband (DNA extracted from newborns’ buccal swabs) using a high stringency GenCall score of 0.75. Using SNPs where the mother and father differed in their homozygous genotypes (mother AA and father BB or vice versa), we determined the presence of both parental chromosomes (genotype AB) in the proband across each chromosome and genomewide. For chromosomal mosaicism, we used the logR and B allele frequencies, which is sensitive >20% for mosaicism detection ^19,20^.

## RESULTS

### Study Participants

A total of 1,603 IVF cycles from 1,190 patients were assessed for eligibility from September 2018 through July 2019 resulting in 41 (2.6%) without fertilization, 225 (14.0%) without blastocyst development, 490 (30.6%) with all aneuploid embryos and 847 (52.8%) with at least one euploid or putative mosaic embryo. In total, 783 patients were enrolled in this trial, leading to 847 single-embryo transfers. Embryo morphology-based embryo selection led to the transfer of 484 uniform euploid embryos (Group A), 282 putative low mosaic embryos (Group B) and 131 putative moderate mosaic embryos (Group C) (**Figure 1**). Baseline characteristics and main IVF cycle outcomes of patients that entered the study are shown in **Table 1**. Main indication for aneuploidy testing was advanced maternal age (AMA, 73.6%) followed by recurrent implantation failure (RIF, 4.1%) and recurrent pregnancy loss (RPL, 3.5%).

**Table 1.**
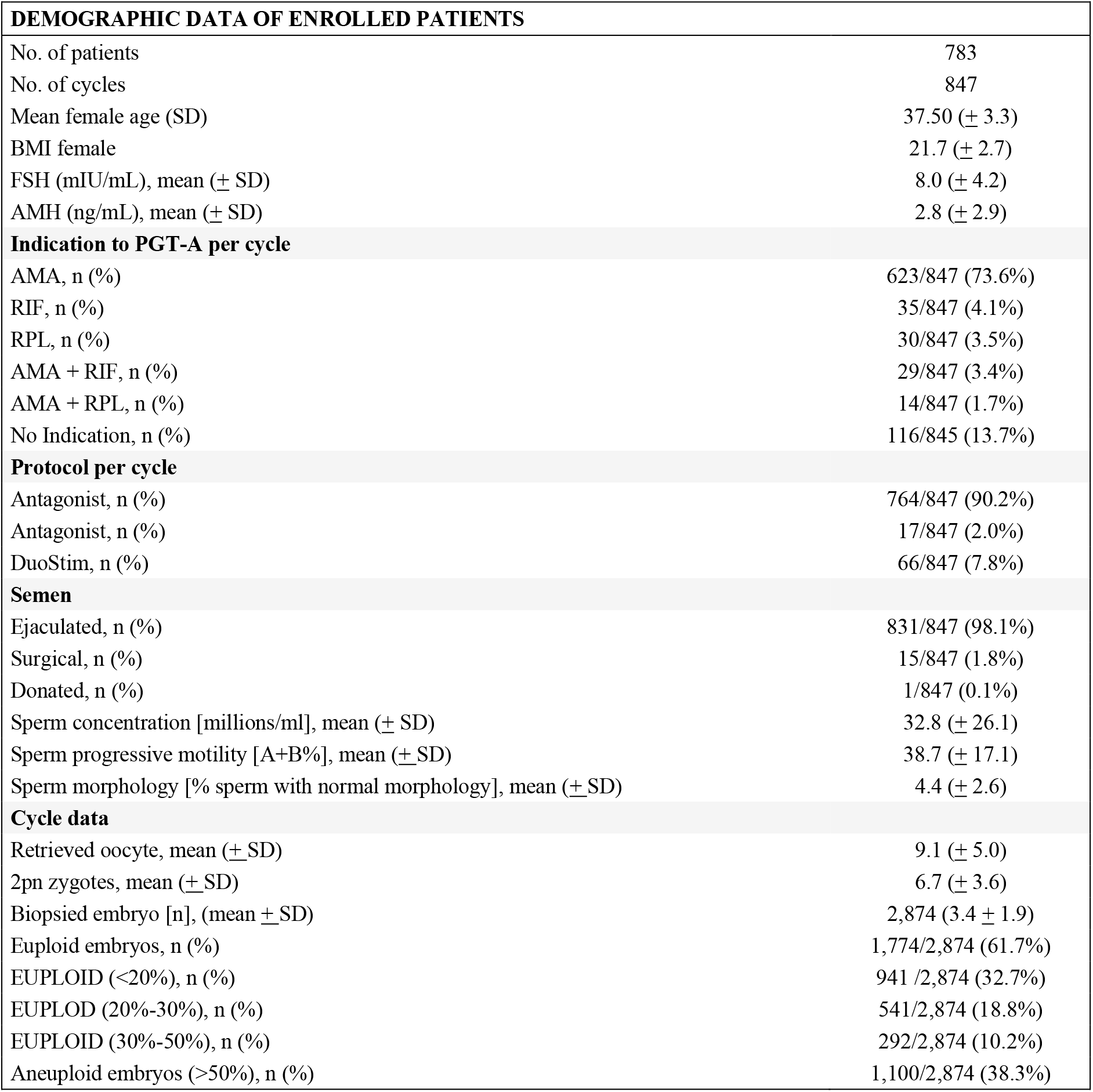
Demographic data of patients enrolled in the trial.

| **DEMOGRAPHIC DATA OF ENROLLED PATIENTS** |  |
| --- | --- |
| No. of patients | 783 |
| No. of cycles | 847 |
| Mean female age (SD) | 37.50 (± 3.3) |
| BMI female | 21.7 (± 2.7) |
| FSH (mIU/mL), mean (± SD) | 8.0 (± 4.2) |
| AMH (ng/mL), mean (± SD) | 2.8 (± 2.9) |
| **Indication to PGT-A per cycle** |  |
| AMA, n (%) | 623/847 (73.6%) |
| RIF, n (%) | 35/847 (4.1%) |
| RPL, n (%) | 30/847 (3.5%) |
| AMA + RIF, n (%) | 29/847 (3.4%) |
| AMA + RPL, n (%) | 14/847 (1.7%) |
| No Indication, n (%) | 116/845 (13.7%) |
| **Protocol per cycle** |  |
| Antagonist, n (%) | 764/847 (90.2%) |
| Antagonist, n (%) | 17/847 (2.0%) |
| DuoStim, n (%) | 66/847 (7.8%) |
| **Semen** |  |
| Ejaculated, n (%) | 831/847 (98.1%) |
| Surgical, n (%) | 15/847 (1.8%) |
| Donated, n (%) | 1/847 (0.1%) |
| Sperm concentration [millions/ml], mean (± SD) | 32.8 (± 26.1) |
| Sperm progressive motility [A+B%], mean (± SD) | 38.7 (± 17.1) |
| Sperm morphology [% sperm with normal morphology], mean (± SD) | 4.4 (± 2.6) |
| **Cycle data** |  |
| Retrieved oocyte, mean (± SD) | 9.1 (± 5.0) |
| 2pn zygotes, mean (± SD) | 6.7 (± 3.6) |
| Biopsied embryo [n], (mean ± SD) | 2,874 (3.4 ± 1.9) |
| Euploid embryos, n (%) | 1,774/2,874 (61.7%) |
| EUPLOID (<20%), n (%) | 941 /2,874 (32.7%) |
| EUPLOID (20%-30%), n (%) | 541/2,874 (18.8%) |
| EUPLOID (30%-50%), n (%) | 292/2,874 (10.2%) |
| Aneuploid embryos (>50%), n (%) | 1,100/2,874 (38.3%) |

**Figure 1.**
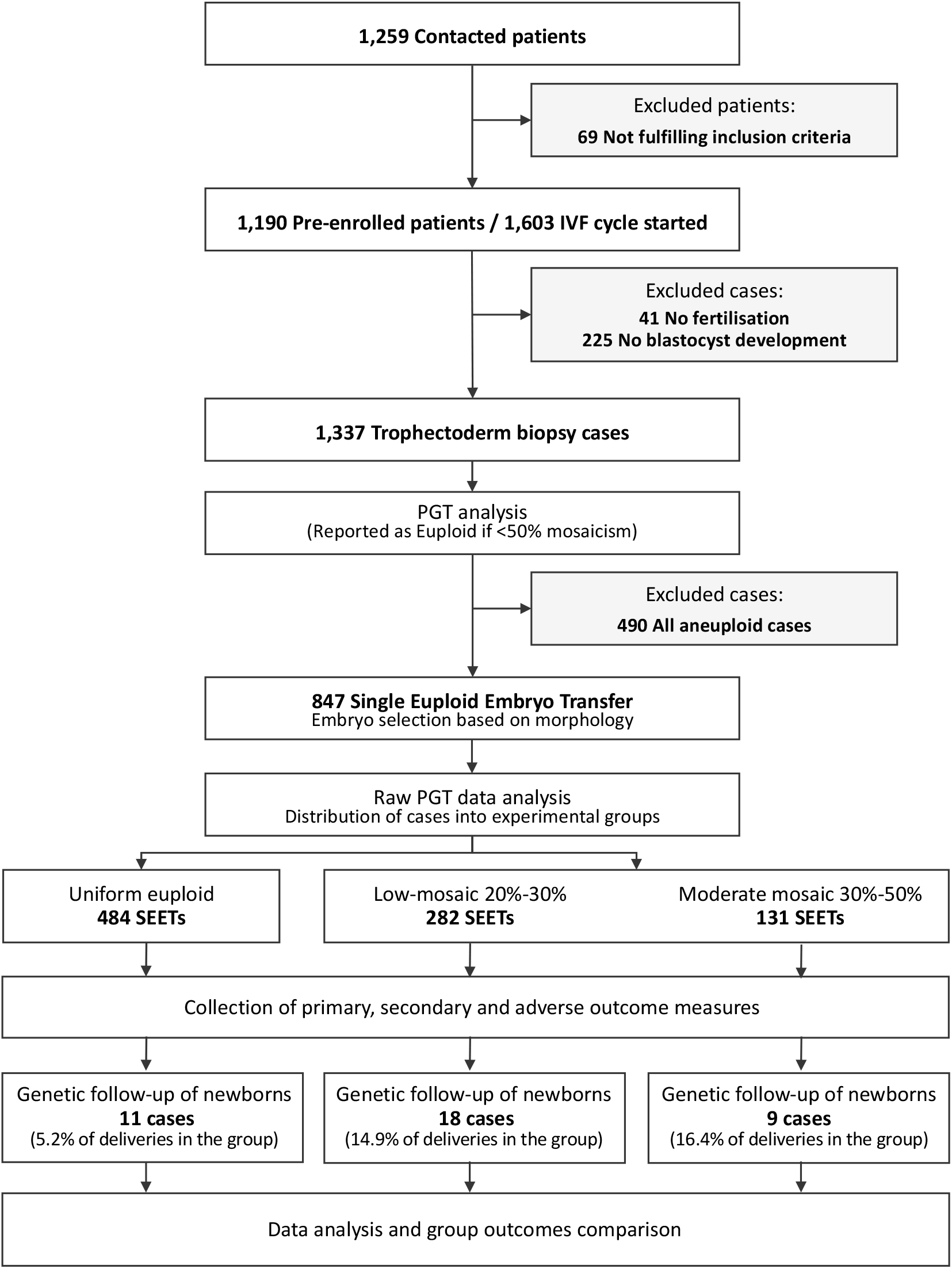
Study Enrollment diagram.

Primary and secondary outcomes could be monitored for all cases, apart for mean gestational age at birth and mean birth weight which were obtained in 97% of cases. A minority of miscarriages could be characterized cytogenetically by POC analysis (n= 4/52; 7.7%) and only 26 pregnancies underwent invasive prenatal diagnosis (n= 26/388, 6.7%) (i.e., CVS and/or amniocentesis). A total of 50 samples were collected from either putative mosaic (n= 36) and uniformly euploid (n= 14) embryo-derived newborns. Of these, 38 passed QC and were selected for molecular testing follow-up involving post-natal karyotyping and genotyping (see **Supplementary Appendix** for further information on enrolment of families in the genotyping follow-up study). All remaining cases from putative mosaic embryos declined or were not available to participate in this phase of the study.

### Primary and secondary outcomes

LB rates derived from euploid embryos, low or moderate-degree mosaic embryos were 43.4% (95% C.I. 38.9 - 47.9), 42.9% (95% C.I. 37.1 - 48.9) and 42% (95% C.I. 33.4 - 50.9), respectively. No difference was detected for the primary outcome between euploid and mosaic low/moderate categories (OR= 0.96; 95% C.I. 0.743 to 1.263; P=0.816). The non-inferiority endpoint was met as the confidence interval for the difference fell below the planned 7.5% margin (95% C.I. −5.7 - 7.3). Also, no statistically significant difference was observed comparing moderate vs low degree mosaic embryos (42.9%, 95% C.I. 37.1 - 48.9 vs 42.0%, 95% C.I. 33.4 - 50.9; P=0.92). There were no significant differences across groups in terms of ongoing pregnancy, clinical pregnancy, biochemical pregnancy, multiple pregnancy, or miscarriage rates (**Table 2**). In terms of perinatal outcomes, the incidence of obstetrical complications, congenital anomalies, and neonatal death were not significantly different across the three groups, as well as the gestational period and birth weight (**Table 2**). Additionally, the number of chromosomes showing a mosaic configuration (commonly referred as complex mosaic) was not associated with any of the primary or secondary outcomes investigated (see **Supplementary Appendix** for additional data and logistic regression analyses, **Table S1 and S2**). In summary, none of the OR and adjusted-OR for euploid vs low/moderate mosaic categories were statistically significant, with 95% C.I. margins fairly close to the unit. At a multivariate analysis level, an effect on LBR was observed for poor quality blastocyst morphology (AOR 0.56 compared to the top-quality category, 95% C.I. 0.35 – 0.89; P=0.0146), day of biopsy (AOR 0.68 per day, 95% C.I. 0.51-0.90, P=0.008) and surgical origin of sperm (AOR 0.158, 95% C.I. 0.04 – 0.75, P=0.020). No significant effects were detected at multivariate analysis for miscarriage and biochemical pregnancy loss.

**Table 2.** Reproductive outcomes after single embryo transfer.

|  | **GROUP A**<br>**EUPLOID** | **GROUP B**<br>**LOW MOSAIC**<br>**(20-30% VARIATION)** | **GROUP C**<br>**MODERATE MOSAIC**<br>**(30-50% VARIATION)** | **ADJ OR**<br>**(95% C.I.**<br>**P-VALUE)** |
| --- | --- | --- | --- | --- |
| **TEST SETS,**<br>**N** | 484 | 282 | 131 |  |
| **POSITIVE**<br>**PREGNANCY TEST,**<br>**% (N)\*** | 55.8%<br>(270/484) | 55.0%<br>(155/282) | 55.7%<br>(73/131) | **0.98**<br>(0.75-1.27; 0.86) |
| **BIOCHEMICAL**<br>**PREGNANCY LOSS,**<br>**% (N)** | 10.7%<br>(29/270) | 12.3%<br>(19/155) | 13.7%<br>(10/73) | **1.18**<br>(0.69-2.02; 0.53) |
| **MISCARRIAGE,**<br>**% (N)** | 12.0%<br>(29/241) | 11.0%<br>(15/136) | 12.7%<br>(8/63) | **0.89**<br>(0.50-1.55; 0.69) |
| **LIVE BIRTH,**<br>**% (N)** | 43.4%<br>(210/484) | 42.9%<br>(121/282) | 42.0%<br>(55/131) | **0.97**<br>(0.74-1.26; 0.82) |
| **MONOCHORIAL**<br>**TWINS DELIVERY, N** | 1 | 1 | 1 |  |
| **GESTATIONAL AGE,**<br>**MEAN (95%C.I.)** | 38.4<br>(38.0-38.7) | 38.2<br>(37.9-38.6) | 38.1<br>(38.0-38.5) |  |
| **BIRTH WEIGHT,**<br>**MEAN (95%C.I.)** | 3,286<br>(3,200-3,371) | 3,174<br>(3,080-3,267) | 3,130<br>(2,950-3,310) |  |

\* Data were imputed in two cases: two patients with no pregnancy-test result were assumed not be pregnant. Biochemical pregnancy is defined by a positive pregnancy test. Implantation rate as the number of gestational sacs observed by vaginal ultrasound at the 5<sup>th</sup> gestational week divided by the number of embryos transferred. Multiple pregnancy is defined by any scan with more than one heartbeat or gestational sac at the stage of clinical pregnancy (approximately 6 weeks). Miscarriages are losses of clinical pregnancy before 20 weeks, excluding ectopic pregnancy. The number of deliveries that resulted in at least one live birth per embryo transfer after 22 weeks of gestation. CI denotes confidence interval. (See outcome definition in the Supplementary appendix).

### Adverse outcomes and cytogenetic follow-up of pregnancies and newborns

Four of the 52 miscarriage cases underwent POC analysis by standard cytogenetic analysis, whilst only 26 of all sustained pregnancies derived (26/388, 6.7%) underwent prenatal diagnosis by amniocentesis (Group A = 15, Group B = 6; Group C = 3) or by CVS as first tier (2 from Group A). Of note, all prenatal diagnoses displayed an euploid karyotype except in one pregnancy from Group A (uniformly euploid) which showed a 20% mosaicism for chromosome 22 during CVS cytogenetic analysis. However, confirmatory amniocentesis failed to support the previous finding identifying euploid karyotypes in all of the 50 metaphases analysed.

Postnatal genotyping of newborns was possible for 38 families (9.8% of all newborns derived from the study) (see **Supplementary Appendix** for recruitment strategy, sample collection and analytical insights). In detail, postnatal genetic analysis was conducted on 5.2%, 14.9% and 16.4% of newborns derived from Group A (n= 11/210), Group B (n= 18/121) and Group C (n= 9/55), respectively. All genotyping tests showed fully normal karyotypes and absence of UPD (**Figure 2** and **Figure S1** in the **Supplementary Appendix**). At birth, no babies showed abnormalities associated with an aberrant karyotype attributable to prenatal mosaicism. One major abnormality was observed among the moderate-grade mosaicism cases (Group C). The baby was born with a diagnosis of Beckwith-Wiedemann Syndrome caused by hypomethylation in the region KvDMR/IC2. The PGT-A profile of the embryo was classified in the moderate-grade mosaicism for a chromosome unrelated to this condition. Because imprinting defects are not diagnostic targets of PGT-A, the NGS analysis did not reveal any indication of UPD at the disease locus.

**Figure 2.**
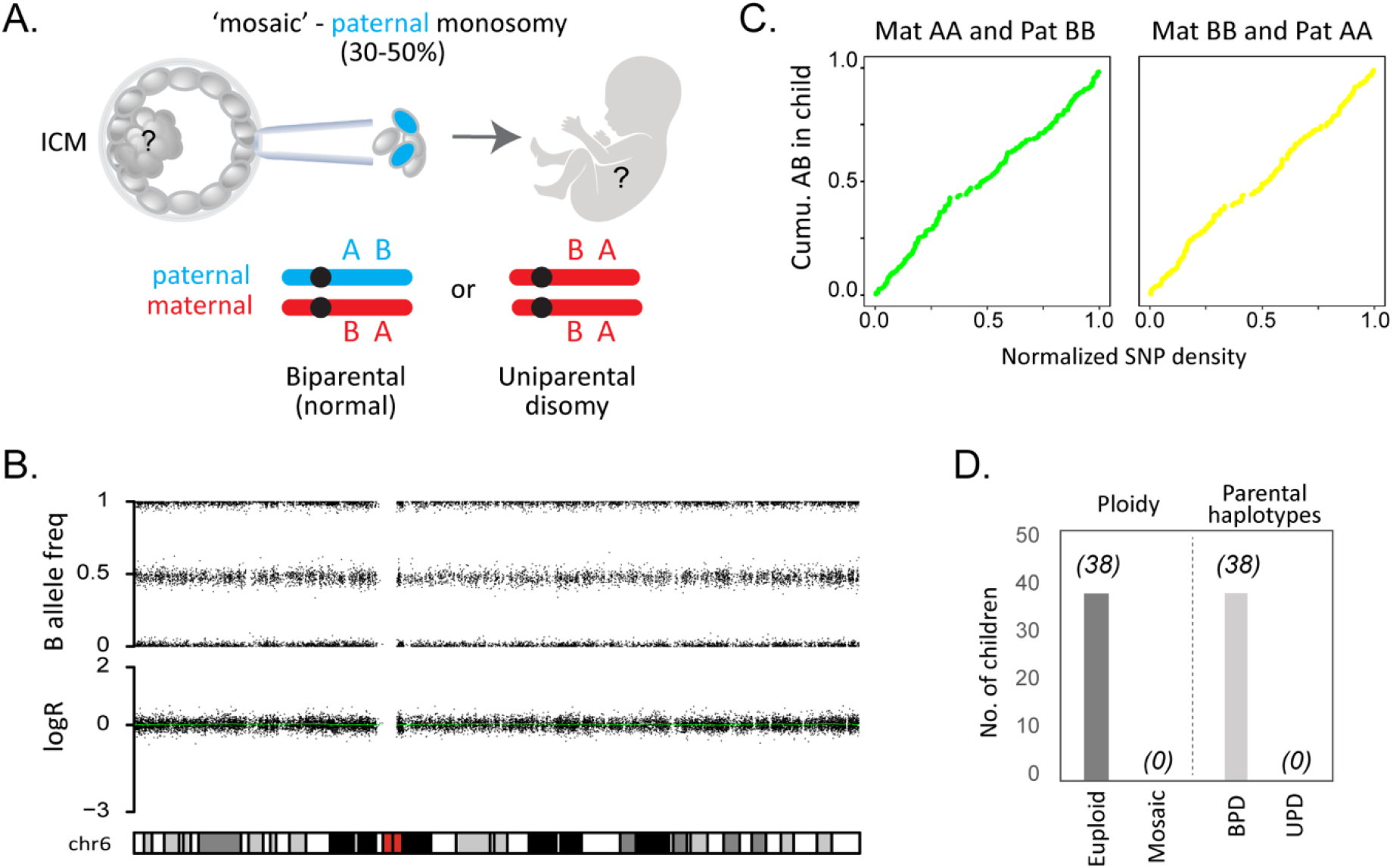
Euploid biparental inheritance in children born from ‘mosaic’ embryo transfer. A) Illustration of a mosaic paternal monosomy inferred from the trophectoderm biopsy. The fetal tissues derive from the inner cell mass that may contain biparental chromosomes uniparental, or a mixture. Supporting SNPs where the maternal and paternal genotypes are homozygous but carry opposite alleles (AA and BB or vice versa) can be used to determine the presence or absence of parental chromosomes. B) LogR and B allele frequencies for chr. 6 from a child born from Group C. C) Cumulative AB genotypes in the child of supporting SNPs across chr. 6. D) Number (No.) of children investigated with post-natal SNPa testing. Total number of samples showing euploid or mosaic karyotype (‘Ploidy’) or containing both parental chromosomes (biparental disomy, BPD) or two homologous chromosomes from the same parent (UPD).

### Probabilistic analysis of treatment efficacy loss

To further challenge the common trend that discourages the transfer of putative mosaic embryos, we employed the data derived from this study to determine the impact of putative mosaicism calling on clinical outcomes. A theoretical model of the impact of low-moderate mosaicism diagnostic calls on cumulative treatment outcomes was produced based on incidence of putative mosaicism and clinical outcome derived from this trial (∼43% LBR). An overall reduction of 24% and 7% was observed if the live births achieved by the use of putative low and moderate mosaic embryos were removed (**Figure 3A**). In an optimistic model using the combined probability of live-birth rate employing all transferrable embryos, an overall relative reduction in cumulative LBR of 11% was expected if all embryos showing moderate-degree mosaicism were excluded from transfer (**Figure 3B**). Based on the same optimistic model, an overall reduction of 36% in cumulative live births following IVF/PGT-A treatment was expected if both embryos showing low and moderate-degree mosaicism were excluded from transfer (**Figure 3B**). Source data are available in **Table S3** in the **Supplementary Appendix**.

**Figure 3.**
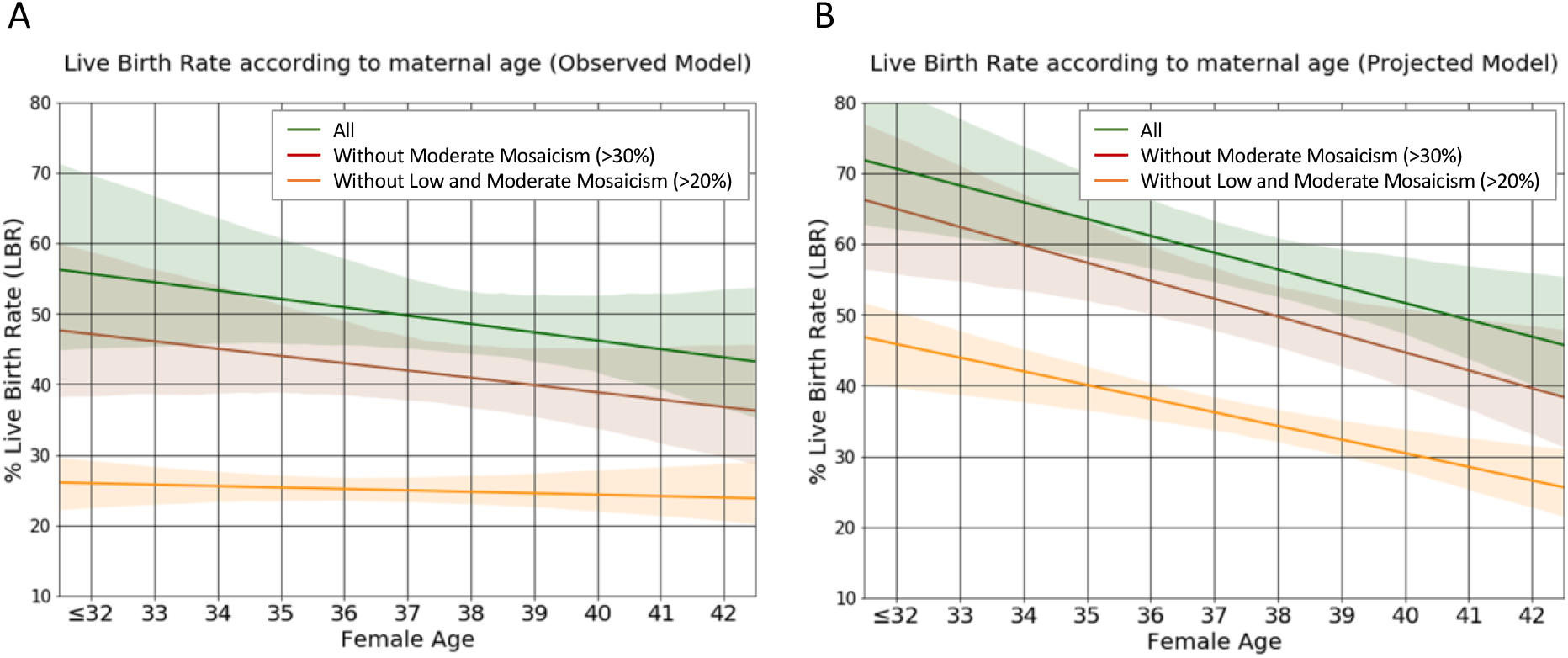
Estimated impact on cumulative live birth rates in case putative mosaicism embryo are excluded from clinical use. Expected cumulative LBR per cycle with utilization of all embryos (euploid + putative mosaics) obtained per each cycle is shown in *green* line across the board of female age. *Brown* line projects the situation where putative moderate mosaic embryos are not utilized (−7% for the observed model and −11% overall relative reduction for the projected model). *Orange* line depicts a scenario where all putative mosaics above 20% variability were excluded from transfer (−24% for the observed model and −36% overall relative reduction for the projected model). A) Observed relative reduction in CLBR per cycle based on actual data from this trial. B) Optimistic model accounting for the combined probability of LBR in the case all transferable embryos are utilized. This modelling is based on the optimistic scenario, assuming that patients with embryos available for transfer who did not already returned for a subsequent replacement cycles would have the same chance of a pregnancy resulting in a live birth as the recorded LBR per embryo transfer in the whole euploid category (i.e., 43%). The mosaicism incidence is plotted based on the rate observed in the trial.

## DISCUSSION

This is the first prospective multicentre, double-blind, non-selection trial aimed at assessing the reproductive potential and safety of low to moderate-degree mosaic embryos by minimizing biases deriving from patients’ prognoses and embryo prioritization based on uncertain PGT-A findings. Identification of embryo mosaicism is an extremely challenging procedure due to both technical and biological limitations of the diagnosis, combined with the heterogeneity of cells biopsied from the trophectoderm, with different biological states of the embryo providing the same analytical profile, the stochastic nature of their sampling, and the marginal signal variability caused by DNA amplification procedures of minimal amount of material.

Past retrospective studies showed lower clinical performance of putative mosaic embryos compared to uniformly euploid ones ^16^. However, they involved the transfer of putative mosaic embryos mainly to patients that had previous failed implantations with uniformly euploid embryos, thus introducing a strong selection bias. As a result of these studies, the transfer of putative mosaic embryos has been abandoned, with reports showing that only 3% are used in clinical treatment ^7^.

By integrating a non-selection design, this trial shows that putative mosaic embryos result in not only comparable clinical outcomes in terms of positive pregnancy, miscarriage and sustained implantation rates but also that newborns derived from embryos diagnosed with putative low/moderate mosaicism are not associated with chromosomal abnormalities at birth. Therefore, the evidence of non-inferior reproductive performance and equivalent safety outcomes of putative mosaic embryos shown by this trial, suggests no clinical utility of reporting mosaicism based on intermediate chromosome copy number deviations up to 50% as currently widely used in PGT-A. These data also support the hypothesis that intermediate chromosome copy number up to 50% may result from technical artefacts arising from WGA processing of minute amounts of embryonic cell.

It should be noted that the results reported in this study were obtained through the analysis of NGS raw data independent from any Igenomix proprietary diagnostic algorithm or chromosome-specific consideration, rather than software-elaborated outputs commonly used in PGT-A laboratories. Since different NGS platforms and associated data analysis tools for PGT may vary across brands and laboratories, this approach appeared to provide a common ground to all PGT laboratories, highly reproducible and independent from specific individual settings. Nevertheless, it is important that each laboratory performs and validates its specific algorithms in prospective non-selection studies similar to the one presented here.

In support of our findings, evidence from clinical predictive values associated with the practice of de-prioritization or deselection of putative mosaic embryos is either absent or confirm the lack of clinical utility. Indeed, positive predictive value of a putative mosaic finding in PGT-A has been confirmed in only one case from over a thousand of allegedly mosaic embryos transferred to date ^21^. Combined with the result from this trial, the risk of having a pregnancy affected by the same mosaicism detected in PGT-A is extremely low based on clinical experience and does not appear to justify current practice of electing invasive prenatal diagnosis by amniocentesis, which has a known iatrogenic risk of abortion of about 0.3% ^8,9^. With regards to negative predictive value, that is the likelihood of reducing the prevalence of true mosaic pregnancies by avoiding the clinical utilization of putative mosaic embryos, data are lacking, and a large sample size would be needed considering the low general prevalence of the condition (around 0.3% of pregnancies). However, few observations so far have highlighted high rates of true mosaicism findings in cytogenetic analysis of POCs from uniformly euploid embryos diagnosed with high resolution aCGH and NGS ^22^. Furthermore, in our trial, the only instance of mosaicism detected in the cytogenetic follow-up of pregnancies and newborns was identified following the transfer a uniformly euploid embryo, further suggesting the inherent limitation in detecting or excluding mosaicism in PGT-A cycle with sufficient accuracy.

The current clinical management of putative mosaic embryos focuses on embryo selection based on the presence of an intermediate chromosome copy number rather than well-established embryo morphological grading. This has the potential not only to drastically reduce the likelihood of pregnancy per cycle, but also to expose patients to increased risk of pregnancy loss by favouring transfer of euploid embryos with poor morphology – known to carry increased risk of resulting in euploid miscarriage – over putative mosaic embryos with better morphological grade ^23–25^. An elaboration of the results of this trial show that excluding putative mosaic embryos (either with low or moderate degree of mosaicism) drastically reduces cumulative pregnancy rate per cycle started (up to −34%) without improving any clinical outcome measure associated with patient’s safety (e.g., miscarriage, chromosomally abnormal conception).

Moreover, clinical management of mosaic embryos demonstrates a range of associated negative consequences, including additional genetic counselling sessions, intensified anxiety and distress, higher costs, increased adoption of invasive prenatal diagnosis, and potential wastage of otherwise normal and healthy pregnancies ^26,27^.

In summary, this trial shows that embryos diagnosed with putative mosaicism produce comparable reproductive outcomes and normal newborn karyotypes to embryos diagnosed as uniformly euploid. The evidence provided by this trial should be taken into consideration by professional societies publishing guidelines on preimplantation genetic testing for aneuploidies. Furthermore, next developments in PGT-A algorithms and aneuploidy classification criteria should benefit from using clinical data from non-selection trials before being incorporating in routine practice.

## Data Availability

All data included in this manuscript are available as supplementary materials or upon authors' request.

